# Perioperative Safety, Surgical Morbidity, and Survival Outcomes of Neoadjuvant Chemotherapy (NACT-IDS) versus Primary Debulking Surgery (PDS) in Elderly Patients with Advanced Epithelial Ovarian Cancer: A Systematic Review and Meta-Analysis

**DOI:** 10.64898/2026.11.12.26367937

**Authors:** Amirhossein Rafiee, Mohammad Ali Boroumand, Shokouh Taghipour Zahi

## Abstract

**Background:** Advanced epithelial ovarian cancer disproportionately affects older women. While primary debulking surgery (PDS) is the traditional gold standard, it carries substantial morbidity and mortality risks in frail, older patients. Neoadjuvant chemotherapy followed by interval debulking surgery (NACT-IDS) can downstage disease and mitigate these surgical risks, but high-level evidence syntheses specific to geriatric populations remain limited.

**Methods:** This systematic review and meta-analysis adhered to PRISMA 2020 guidelines and was registered on PROSPERO (CRD420261461644). Five electronic databases and ClinicalTrials.gov were searched for comparative studies evaluating NACT-IDS versus PDS in women aged 70 years (or 65 years in defined geriatric cohorts) with FIGO Stage IIIC/IV epithelial ovarian cancer. Outcomes were synthesized using random-effects models. Methodological quality was assessed via the ROBINS-I tool, cumulative evidence reliability via Trial Sequential Analysis (TSA), and overall evidence certainty via the GRADE framework.

**Results:** Eleven retrospective cohort studies encompassing over 16,500 older patients met the inclusion criteria. NACT-IDS yielded an 80% reduction in unadjusted 30-day perioperative mortality compared to PDS (RR = 0.20, 95% CI [0.05, 0.89], p = 0.034). Furthermore, NACT-IDS significantly decreased hospital length of stay (MD = -3.12 days, p = 0.0044) and increased the likelihood of complete macroscopic resection (R0 = 0 cm) by 74% (RR = 1.74, 95% CI [1.01, 3.02], p = 0.047). Severe complications (Clavien-Dindo 3) trended lower with NACT-IDS (RR = 0.38, 95% CI [0.13, 1.08]). Unadjusted overall survival models demonstrated a 13% increased hazard of death for NACT-IDS (HR = 1.13, 95% CI [1.04, 1.23]); however, this primarily reflects profound selection bias (confounding by indication) where the frailest patients were systematically triaged to the NACT-IDS pathway. The overall certainty of evidence was appraised as Very Low.

**Conclusion:** For older women with advanced epithelial ovarian cancer, NACT-IDS significantly improves perioperative safety, accelerates hospital recovery, and maximizes complete resection rates compared to upfront PDS. While retrospective survival data are heavily skewed by channeling bias, NACT-IDS effectively protects physiological reserve and should be strongly considered as the standard initial management strategy for this vulnerable demographic.

## Introduction

### Epidemiological Context & Surgical Vulnerability in Older Adults

Epithelial ovarian cancer is primarily a disease of older women (1). Patients aged 65 and older account for 50% of all diagnoses (2). Nearly half of the affected population exceeds 64 years of age, and 25% surpass 74 years (3). Incidence peaks between 75 and 79 years (1). This demographic bears a disproportionate disease burden, representing 40–45% of incident cases and 65% of disease-specific mortality among women over 65 (4). Incidence and mortality rates for women aged 50 and older reach 27.6 and 17.6 per 100,000, respectively (5). Conversely, women under 50 experience an incidence of 6.0 per 100,000 and a mortality rate of 2.7 per 100,000 (5). Age dictates survival. United Kingdom data demonstrate a 10-year survival rate of just 21.5% for patients over 75, compared to 81.2% for those aged 15 to 44 (1). Demographic projections indicate the population of patients over 65 will double within two decades (2).

Aging manifests as a progressive deterioration of stress tolerance, driven by diminished functional reserve across multiple organ systems (2). The cardiovascular, renal, pulmonary, central nervous, and skeletomuscular systems exhibit steady depletion, fundamentally altering the physiological response to acute stressors (6). These age-related functional deficits elevate the risk of adverse surgical outcomes relative to younger cohorts (7). Altered baseline physiology directly modifies the pharmacokinetics and pharmacodynamics of systemic medications, particularly anesthetic agents (6).

Frailty represents a distinct clinical syndrome defined by depleted homeostatic reserve and heightened vulnerability to external stressors (4,8,9). The compounding stressors of ovarian cancer and aggressive multimodality therapy frequently precipitate a rapid transition from independent self-sufficiency to severe disability in frail older women (9). Preoperative frailty severely complicates surgical intervention (10). During extensive cytoreductive surgery for ovarian cancer, frailty consistently predicts elevated postoperative morbidity, extended hospital stays, and compromised survival (11). Meta-analytic data confirm frailty significantly increases the risk of all complications (OR: 1.61) and major complications (OR: 1.80). Frail patients also face a higher probability of readmission (OR: 1.43) (10). Age directly amplifies surgical risk. Compared to patients under 60, the odds ratio for 30-day postoperative mortality escalates to 3.7 for ages 60– 69, 3.1 for ages 70–79, and 9.3 for those 80 and older. Fatal postoperative complications occur in 9% of octogenarians, compared to only 1% of patients under 60. Patients exceeding 70 years, and especially those over 80, demonstrate a higher frequency of postoperative chronic kidney failure, cardiorespiratory events, and neurological deficits (6).

### The Surgical Paradigm: Historical PDS vs. Landmark NACT-IDS Trials

Several landmark randomized controlled trials confirm the non-inferiority of neoadjuvant chemotherapy (NACT) followed by interval debulking surgery versus primary debulking surgery (PDS). The EORTC 55971 trial reported an overall survival of 27.6 months for PDS and 26.9 months for NACT (HR 0.97, p=0.586). Progression-free survival reached 11.6 months and 11.1 months, respectively (HR 0.98, p=0.688) (12). The CHORUS trial evaluated women with stage III or IV high-burden ovarian cancer, establishing that primary chemotherapy yielded non-inferior survival compared to primary surgery. Median overall survival in CHORUS was 22.6 months for primary surgery and 24.1 months for primary chemotherapy (13). The JCOG0602 trial demonstrated an overall survival of 49.0 months with PDS versus 44.3 months with NACT (HR 1.052, p=0.24) (14). The SCORPION trial recorded a median progression-free survival of 15 months for PDS and 14 months for NACT (HR 1.05, p=0.73), alongside a median overall survival of 41 months versus 43 months (HR 1.12, p=0.56) (15).

NACT significantly mitigates surgical morbidity and mortality (13). EORTC 55971 recorded a postoperative mortality rate of 2.5% in the PDS arm versus 0.7% in the NACT arm (16). In the CHORUS trial, grade 3 or 4 postoperative adverse events occurred in 24% of the primary-surgery group compared to 14% of the primary-chemotherapy group (p=0.0007), while deaths within 28 days reached 6% versus <1% (p=0.001) (13). JCOG0602 confirmed a significantly lower incidence of grade 3 or 4 postoperative adverse events following NACT (p=0.003) (14). Intraoperative metrics heavily favor NACT, with median ascitic fluid aspiration and blood loss substantially reduced compared to PDS (2820 mL vs 0 mL, p<0.001) (17). The SCORPION trial localized major postoperative complications primarily to the PDS arm (25.9% vs 7.6%; p=0.0001) (15). Postoperative mortality in SCORPION was 8.3% for PDS, with zero postoperative deaths recorded in the NACT cohort (16). SCORPION demonstrated superior complete resection (R0) rates in the NACT arm, achieving 77.0% compared to 47.6% for PDS (p=0.001) (15). Hospital recovery accelerates under the NACT paradigm. In CHORUS, 93% of primary-chemotherapy patients achieved discharge within 14 days of surgery, compared to 80% of primary-surgery patients (13).

### The Geriatric Knowledge Gap & Clinical Rationale

Older patients with ovarian cancer remain systematically underrepresented in clinical trials (1). This recruitment deficit is a long-standing structural flaw in oncologic research (18). A Southwest Oncology Group (SWOG) analysis of 16,396 patients across 164 trials from the 1990s revealed that patients over 65 comprised only 30% of the study population. A recent Surveillance, Epidemiology, and End Results (SEER) survey found that a mere 9% of cancer patients older than 75 enrolled in novel therapeutic trials (3). Women exceeding 70 years of age were entirely excluded from these studies (18). First-line phase 3 trials for advanced ovarian cancer consistently selected young cohorts. Median ages were 59 years in MITO 7-ENGOT-Ov-10, 57 years in ICON7, 62 years in EORTC 55971, and 57 years in the Gynecological Oncology Group study. Only the CHORUS trial successfully recruited a higher average age demographic (13). Historically, investigators excluded older women from aggressive surgical trials based on inaccurate underestimations of life expectancy beyond age 70 and presumed physiological intolerance to radical treatment (19).

Standard management protocols rely on data derived from younger populations. When older patients do enter major trials, they represent a highly selected, exceptionally fit subgroup that fails to reflect the true geriatric population (20). Consequently, methodologically rigorous conclusions drawn from younger cohorts do not seamlessly translate to older adults (18). Aging drives a progressive loss of stress tolerance, rooted in the depletion of functional organ reserve and the erosion of personal and social resources (2). The cardiovascular, renal, pulmonary, central nervous, and skeletomuscular systems lose capacity over time, fundamentally altering physiological responses to stress (6). Renal function can decline by 40% by age 70. This drop in glomerular filtration rate directly amplifies the toxicity of renally excreted agents like cisplatin (3). Altered geriatric physiology shifts the pharmacokinetics and pharmacodynamics of all systemic medications, particularly anesthetics (6). While these age-related physiological changes may remain clinically silent under homeostatic conditions, acute surgical or cytotoxic stress rapidly exposes the underlying lack of functional reserve (9).

Frailty operates as a complex biological syndrome defined by exhausted physiological reserve and an inability to withstand stressors, leaving the patient highly vulnerable to adverse outcomes (4). Preoperative frailty complicates surgical intervention, driving increased complication rates, frequent readmissions, and elevated early and late mortality. Meta-analytic data in ovarian cancer demonstrate that frailty significantly increases the risk of all complications (OR: 1.61) and major complications (OR: 1.80) (10). Failure to identify frailty preoperatively leads to inappropriately aggressive interventions. This oversight yields poor oncologic outcomes, elevated mortality, degraded quality of life, and excessive financial costs (18). Current evidence confirms chronological age alone fails to predict chemotherapy tolerance (4). Future clinical trials must abandon the exclusive focus on younger, fit cohorts and the healthiest subset of older women (3).

### Objectives

The primary objective of this systematic review and meta-analysis is to definitively evaluate the perioperative safety, surgical morbidity, and oncological outcomes of NACT-IDS compared to PDS, specifically in older women (age ≥70 years) with advanced (FIGO Stage IIIC-IV) epithelial ovarian cancer. We hypothesize that NACT-IDS mitigates the physiological insult of radical cytoreduction, thereby reducing severe postoperative complications and short-term mortality in this vulnerable demographic.

## METHODS

### Protocol Registration & PRISMA Statement

This systematic review and meta-analysis was conducted in rigorous adherence to the Preferred Reporting Items for Systematic Reviews and Meta-Analyses (PRISMA) 2020 guidelines. The predefined methodology was prospectively registered in the PROSPERO International Register of Systematic Reviews (Registration No. CRD420261461644).

### Eligibility Criteria & PICO Framework

The Population, Intervention, Comparator, and Outcomes (PICO) framework was utilized to define inclusion criteria. Studies were eligible if they included: (P) female patients designated as elderly (defined as chronological age 70 years, or 65 years if explicitly analyzed as a distinct geriatric cohort) with advanced-stage (FIGO IIIC/IV) epithelial ovarian, fallopian tube, or primary peritoneal carcinoma; (I) treatment with neoadjuvant chemotherapy followed by interval debulking surgery (NACT-IDS); (C) comparison to primary debulking surgery (PDS); and (O) reported on at least one primary safety outcome (perioperative mortality, severe Clavien-Dindo 3 complications), surgical efficacy outcome (complete macroscopic resection, R0 = 0 cm), or survival metric (Overall Survival [OS]). Exclusion criteria encompassed non-comparative single-arm studies, case reports, cohorts lacking isolated geriatric data, and studies analyzing non-epithelial histologies.

### Information Sources & Search Strategy

A comprehensive, systematic literature search was executed without language restrictions across five electronic databases: PubMed/MEDLINE, Embase, Scopus, the Cochrane Central Register of Controlled Trials (CENTRAL), and Google Scholar. To identify unpublished or ongoing trials, ClinicalTrials.gov was also queried. The search strategy utilized a combination of Medical Subject Headings (MeSH) and free-text keywords encompassing the population ("Elderly", "Geriatric", "Aged"), intervention ("Neoadjuvant Therapy", "NACT", "Interval Debulking"), comparator ("Primary Debulking", "PDS", "Cytoreductive Surgery"), and disease state ("Ovarian Neoplasms", "Epithelial Ovarian Cancer").

### Study Selection & Data Extraction

Retrieved citations were imported into EndNote for deduplication and subsequently transferred to the Rayyan web application for screening. Two investigators independently screened titles and abstracts, followed by full-text evaluation of potentially eligible records. Discrepancies were resolved via consensus or consultation with a third senior investigator. Data extraction was performed independently using standardized collection forms, capturing study parameters, patient demographics, surgical variables, and clinical outcomes.

### Methodological Quality & Risk of Bias

Because the final included literature consisted entirely of non-randomized comparative cohort studies, methodological quality was assessed using the Risk Of Bias In Non-randomized Studies - of Interventions (ROBINS-I) tool. Studies were evaluated across seven domains: confounding, selection of participants, classification of interventions, deviations from intended interventions, missing data, measurement of outcomes, and selection of the reported result. Visual summaries of bias risk were generated.

### Statistical Synthesis

Meta-analytic synthesis was performed using the meta package in R (R Foundation for Statistical Computing). Given the anticipated clinical heterogeneity inherent in global surgical practices and geriatric definitions, random-effects models were utilized universally. Statistical heterogeneity was quantified using the statistic and Cochrane’s Q test, with values 75% considered high heterogeneity.

- **Binary Outcomes:** Pooled as Relative Risks (RR) with 95% Confidence Intervals (CI) utilizing the Mantel-Haenszel method. For rare safety endpoints (e.g., perioperative mortality), in accordance with Cochrane methodological guidelines, the routine 0.5 continuity correction was omitted for "double-zero" studies in the primary analysis to prevent the artificial dilution of the effect size toward the null. Penalized models incorporating the continuity correction were executed as sensitivity analyses.
- **Continuous Outcomes:** Variables such as hospital length of stay (days) were synthesized as Mean Differences (MD) using the generic inverse-variance method and the Restricted Maximum-Likelihood (REML) estimator for between-study variance (υ^2^). Imputation of standard deviations from medians and ranges was performed when necessary for sensitivity testing.
- **Survival Outcomes:** Time-to-event data (OS) were pooled as Hazard Ratios (HR) using the generic inverse-variance method.

### Trial Sequential Analysis (TSA)

To control for the risks of Type I (false positive) and Type II (false negative) errors inherent in cumulative meta-analyses, and to assess whether the accrued geriatric sample size was sufficient, Trial Sequential Analysis (TSA) was performed. The Diversity-Adjusted Required Information Size (DARIS) was calculated using an alpha of 5% and a beta of 20% (80% power). O’Brien-Fleming alpha-spending monitoring boundaries were constructed to determine if the cumulative Z-curve crossed into areas of definitive clinical benefit or harm prior to reaching the required information size.

### Certainty of Evidence Assessment

The overall certainty of the body of evidence was appraised using the Grading of Recommendations Assessment, Development and Evaluation (GRADE) framework. Outcomes were graded as High, Moderate, Low, or Very Low certainty based on the observational nature of the starting evidence, combined with downgrades for serious risk of bias (confounding), inconsistency (heterogeneity), indirectness, and imprecision (evaluated via TSA thresholds).

## RESULTS

### Study Selection Flow & Search Yield

The initial literature search yielded 4,691 records. We removed 1,731 duplicates and screened the remaining 2,960 records by abstract. Investigators sought 173 full-text articles for retrieval and fully assessed 161 for eligibility. We excluded 150 articles, primarily due to a lack of separate elderly data (n = 123) or the absence of a direct comparison (n = 20). The final quantitative synthesis included 11 retrospective cohort studies comprising over 16,500 elderly patients (Figure 1).

**Figure 1:**
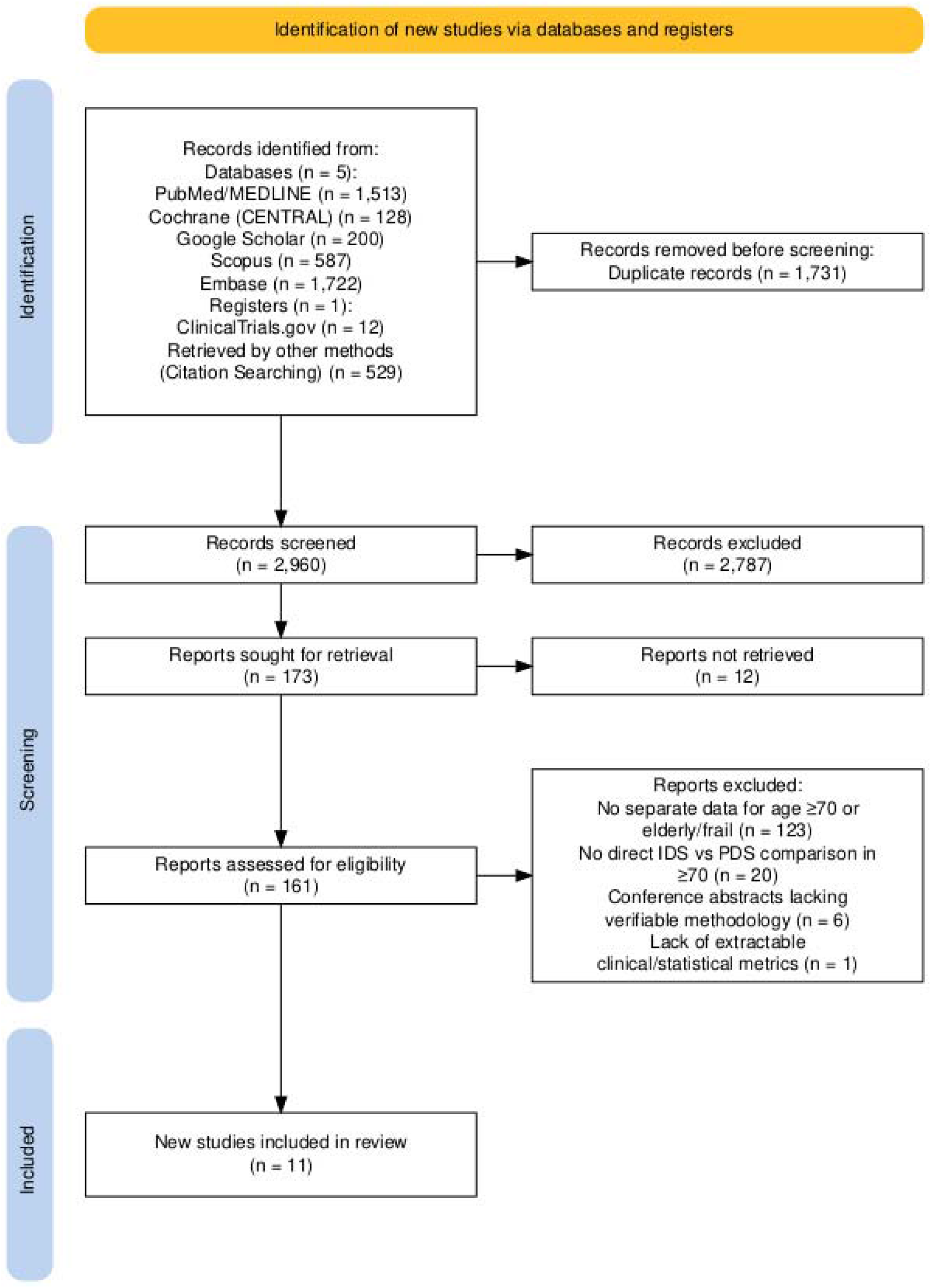
Prisma Flowchart.

### Characteristics of Included Studies

The 11 included cohorts spanned treatment periods from 1991 to 2018. These studies originated across the USA, Canada, Italy, and Denmark. Age cutoffs defining the elderly cohort ranged from ≥65 to ≥75 years (Table 1).

**Table 1:** Baseline Characteristics of Included Studies.

| **Study (Author, Year)** | **Setting & Study Period** | **Age Cutoff** | **Total N (NACT n / PDS n)** | **Median Age** | **FIGO Stage Distribution** | **Chemotherapy/Histology** |
|---|---|---|---|---|---|---|
| Ekmann-Gade, 2023(21) | Denmark, Nationwide (Danish Gynecological Cancer Database & National Patient Registry) (2013–2018) | ≥70 | 417 (235 / 182) | Overall: NR<br>NACT: 74 [72, 76]<br>PDS: 74 [72, 78] | IIIC: 245 (60%), IV: 160 (40%) | **Histology:** NACT: 155 (66%), PDS: 138 (76%)<br>**Chemotherapy:** NR |
| Fanfani, 2012(22) | Italy, Catholic University of Rome and Campobasso (1994–2009) | ≥75 | 32 (10 / 22) | Overall: NR [75-85]<br>NACT: NR<br>PDS: NR | IIIC: 30 (93.7%), IV: 2 (6.3%) | **Histology:** Serous: 24 (75.0%)<br>**Chemotherapy:** Platinum and paclitaxel (6 cycles) |
| Filippova, 2021(23) | USA, Memorial Sloan Kettering Cancer Center (2001–2017) | ≥70 | 430 (199 / 231) | Overall: 75 (72–79)<br>NACT: NR<br>PDS: NR | IIIC: 256 (60%), IV: 152 (35%) | **Histology:** High-Grade Serous: 359 (83%)<br>**Chemotherapy:** Platinum-based (Median 5 cycles) |
| Glasgow, 2013(24) | USA, Yale-New Haven Hospital (1996–2009) | ≥70 | 104 (42 / 62) | Overall: NR<br>NACT: Mean 76.9 (70–89)<br>PDS: Mean 75.9 (70–89) | NACT: IIIC 18 (42.9%) / IV 24 (57.1%)<br>PDS: IIIC 44 (71.0%) / IV 18 (29.0%) | **Histology:** Serous: NACT 31 (73.8%), PDS 33 (53.2%)<br>**Chemotherapy:** Carboplatin and paclitaxel (Median 6 cycles) |
| Matsuo, 2021(25) | USA, SEER Program (2010–2016) | ≥65 | NR (IPTW Cohort) | Overall: NR<br>NACT: NR<br>PDS: NR | NR | **Histology:** High-Grade Serous: 100%<br>**Chemotherapy:** NR |
| McLean, 2010(26) | USA, University of Washington Medical Center (1997–2007) | ≥65 | 175 (34 / 141) | Overall: Mean 72<br>NACT: NR<br>PDS: NR | III: 121 (69.1%), IV: 30 (17.1%) | **Histology:** Papillary Serous: 73% (Age 65-79), 81% (Age ≥80)<br>**Chemotherapy:** IV Carboplatin and paclitaxel (Median 6 cycles) |
| Meyer, 2018(27) | USA, SEER-Medicare database (2000–2013) | ≥66 | 5417 (1221 / 4196) | Overall: NR<br>NACT: Mean 74.1 ± 5.6<br>PDS: Mean 74.1 ± 5.5 | NACT: III 662 (54.2%) / IV 559 (45.8%)<br>PDS: III 2967 (70.7%) / IV 1229 (29.3%) | **Histology:** Serous: NACT 856 (70.1%), PDS 3180 (75.8%)<br>**Chemotherapy:** Platinum/taxane-based |
| Piedimonte, 2020(28) | Canada, Jewish General Hospital, McGill University (2008–2017) | ≥75 | 71 (58 / 13) | Overall: NR<br>NACT: 79 (75–91)<br>PDS: 79 (75–83) | NACT: III 49 (84.5%) / IV 9 (15.5%)<br>PDS: III 11 (84.6%) / IV 2 (15.4%) | **Histology:** High-grade serous and endometrioid<br>**Chemotherapy:** Platinum agent |
| Piedimonte, 2022(29) | Canada, Princess Margaret Cancer Centre (2010–2018) | ≥70 | 168 (99 / 69) | Overall: NR<br>NACT: 75.5 (70–86)<br>PDS: 74 (70–90) | NACT: III 80 (80.8%) / IV 19 (19.2%)<br>PDS: III 63 (91.3%) / IV 6 (8.7%) | **Histology:** High-Grade Serous: NACT 95 (96%), PDS 54 (78.2%)<br>**Chemotherapy:** Carboplatin and paclitaxel (Median 6-7 cycles) |
| Worley, 2013(30) | USA, Brigham and Women's Hospital & Massachusetts General Hospital (2000–2010) | ≥70 | 165 (40 / 125) | Overall: 75<br>NACT: 74 (70–84)<br>PDS: 75 (70–95) | NACT: IIIC 23 (57.5%) / IV 17 (42.5%)<br>PDS: IIIC 108 (86.4%) / IV 17 (13.6%) | **Histology:** Serous: NACT 35 (87.5%), PDS 104 (83.2%)<br>**Chemotherapy:** Carboplatin and paclitaxel (6 cycles) |
| Wright, 2014(31) | USA, SEER-Medicare database (1991–2007) | ≥65 | 7583 (2238 / 5345) | Overall: NR<br>NACT: NR<br>PDS: NR | NACT: III 39.4% / IV 56.7%<br>PDS: III 54.8% / IV 34.2% | **Histology:** Serous: NACT 34.3%, PDS 66.7%<br>**Chemotherapy:** Cytotoxic chemotherapy |

*Abbreviations:* **FIGO**, International Federation of Gynecology and Obstetrics; **IPTW,** Inverse Probability of Treatment Weighting; **IV**, intravenous; **N**, total number of patients; **n**, number of patients in subgroup; **NACT**, neoadjuvant chemotherapy; **NR**, not reported; **PDS**, primary debulking surgery; **SEER**, Surveillance, Epidemiology, and End Results.

### Methodological Quality & Risk of Bias

We assessed methodological quality using the ROBINS-I tool. The overall risk of bias was Serious in 9 studies (81.8%) and Moderate in 2 studies (18.2%). Confounding (D1) represented the most common Serious domain, affecting 72.7% of the included cohorts. No studies demonstrated a Critical risk of bias (Figure 2.1 and Figure 2.2).

**Figure 2.1:**
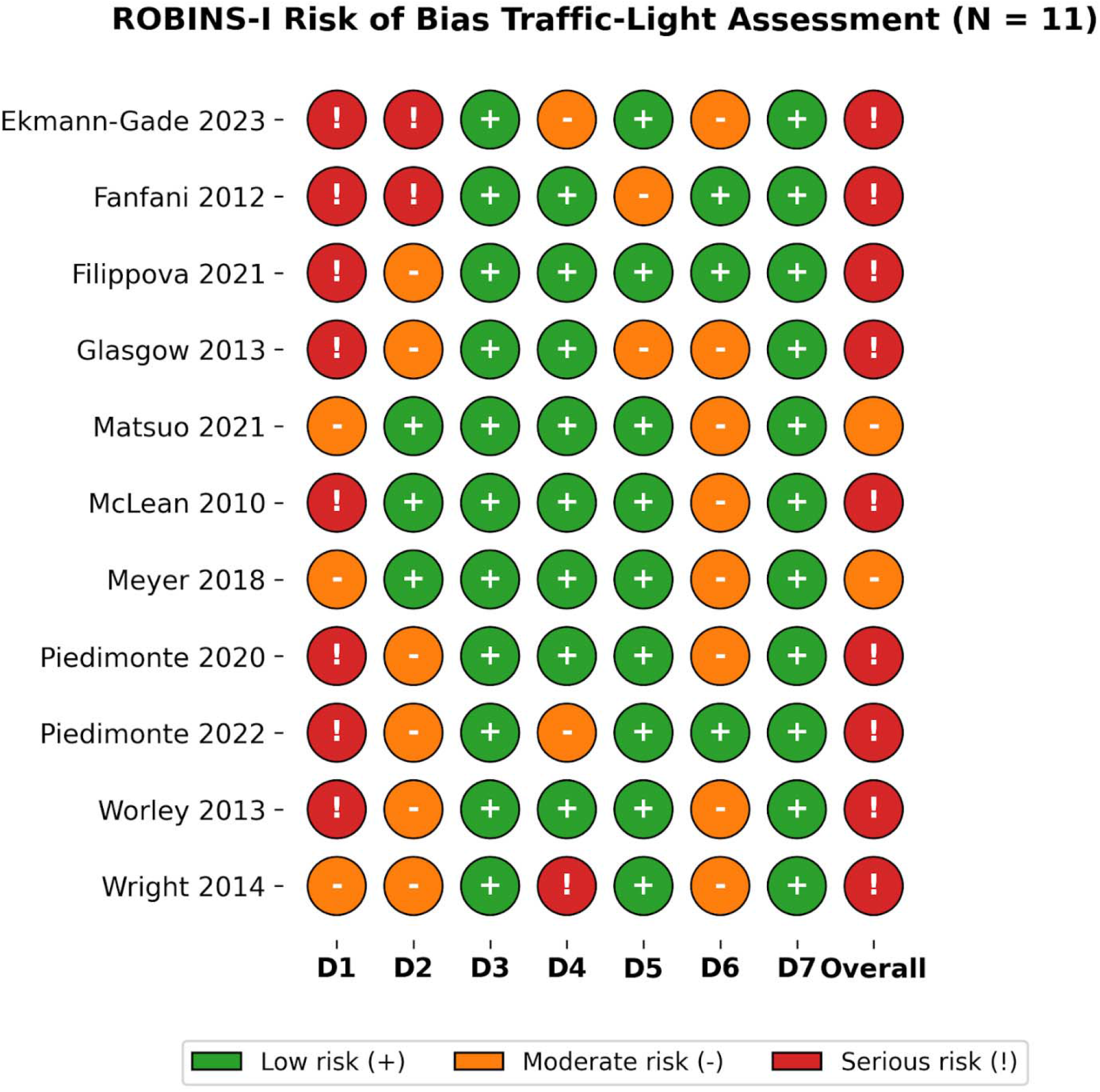
ROBINS-I (Traffic Light View)

**Figure 2.2:**
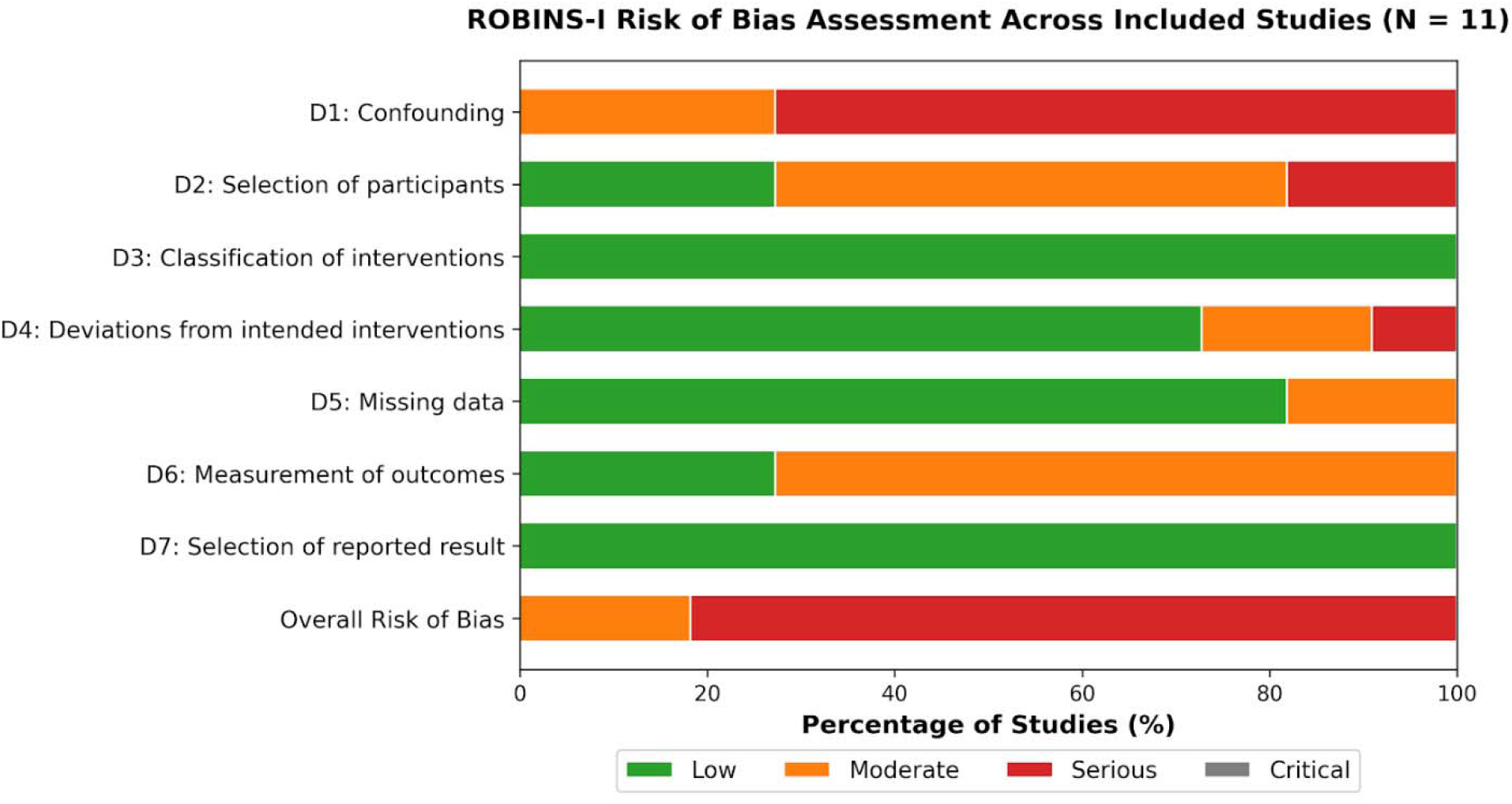
ROBINS-I Summary Bar.

### Primary Binary Safety & Surgical Completeness Endpoints

#### Perioperative Mortality (30-day & broad)

NACT-IDS was associated with an 80% reduction in 30-day perioperative mortality before double-zero continuity correction (RR = 0.20, 95% CI [0.05, 0.89], p = 0.034). Following continuity correction, this protective trend remained but lost statistical significance (RR = 0.27, 95% CI [0.07, 1.09], p = 0.066). Additional details for the continuity corrected version are provided in Supplementary Figures 3.1 and 3.2. Statistical heterogeneity for this outcome was zero (I2 = 0%) (Figure 3.1 & 3.2).

**Figure 3.1:**
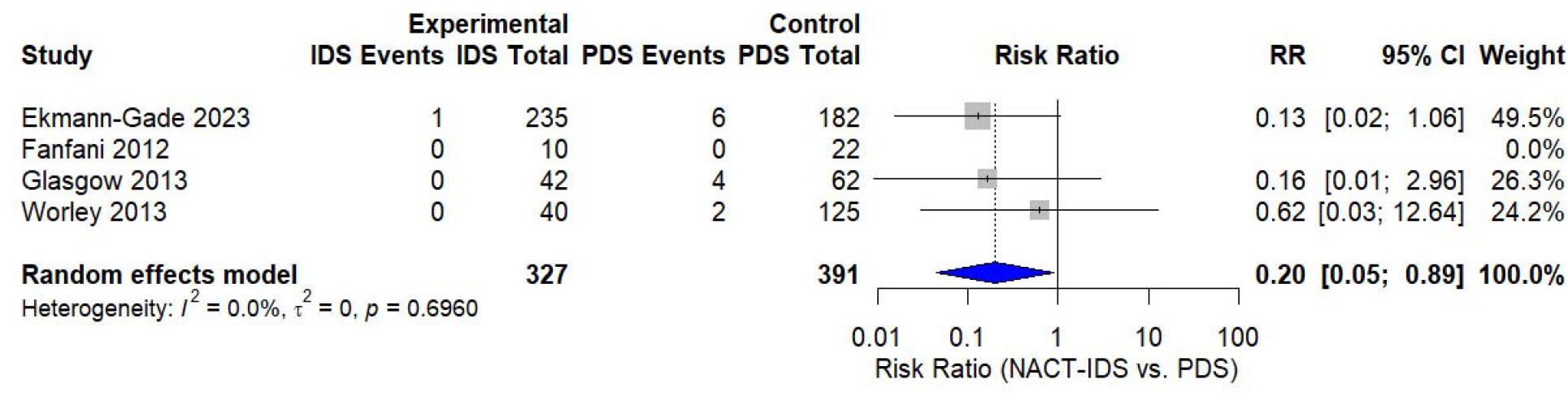
Primary 30-Day Perioperative Mortality.

**Figure 3.2:**
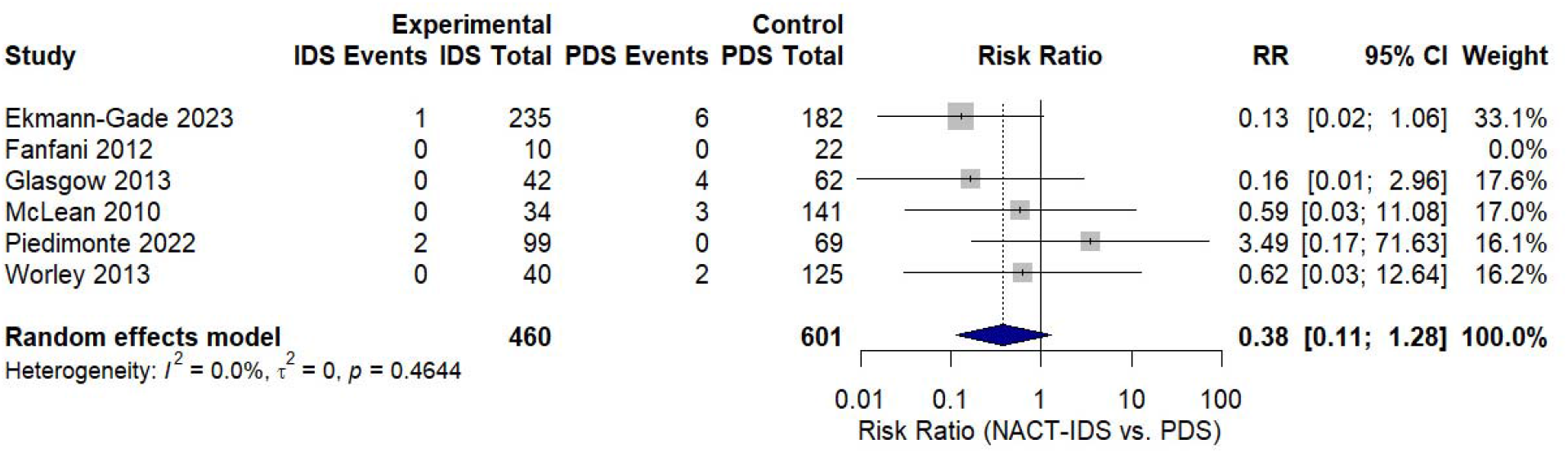
Sensitivity Broad Perioperative Mortality.

**Figure 3.3:**
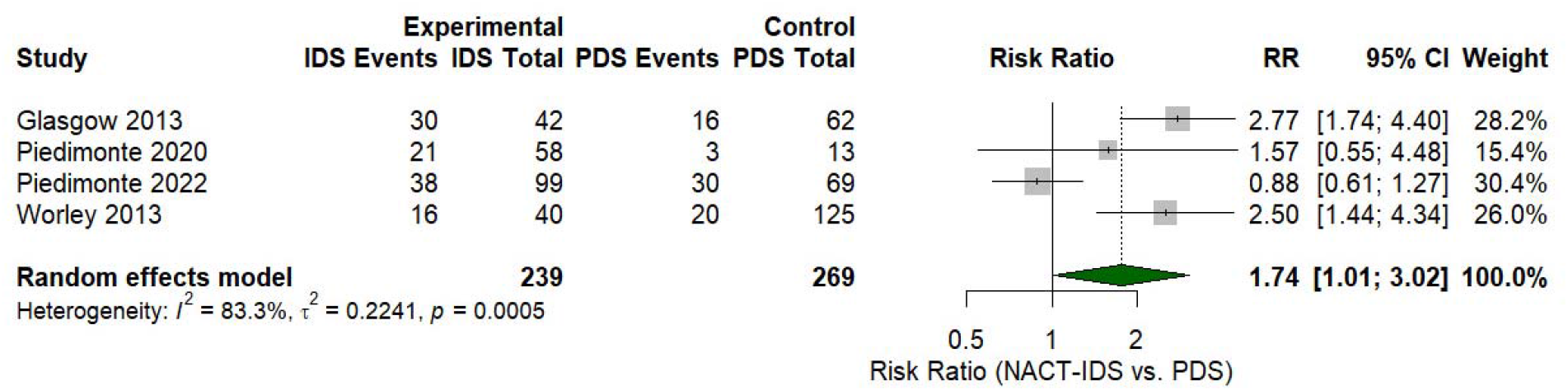
Complete Resection Rate.

**Figure 3.4:**
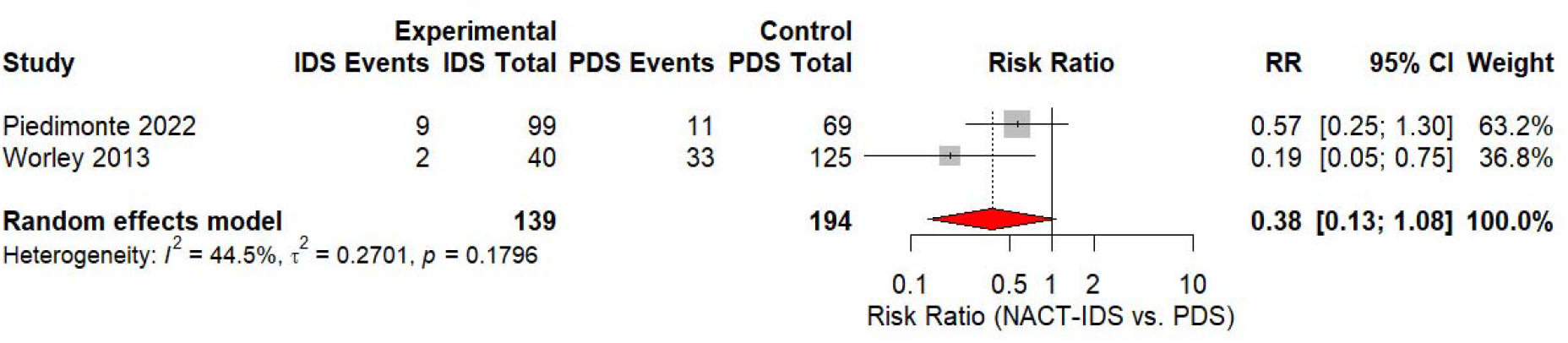
Severe Postoperative Morbidity.

#### Macroscopic Complete Resection (R0 = 0 cm)

NACT-IDS significantly increased the likelihood of complete macroscopic resection (R0 = 0 cm) by 74% compared to PDS (RR = 1.74, 95% CI [1.01, 3.02], p = 0.0476, I2 = 83.3%) (Figure 3.3).

#### Severe Postoperative Morbidity (Clavien-Dindo ≥3)

Analysis of severe complications (Clavien-Dindo ≥3) demonstrated a strong protective trend for NACT-IDS, yielding a 62% reduction (RR = 0.38, 95% CI [0.13, 1.08], p = 0.0687) (Figure 3.4).

### Inpatient Surgical Morbidity: Hospital Length of Stay

NACT-IDS significantly reduced hospital length of stay by a pooled mean difference of - 3.12 days (95% CI [-5.27, -0.97], p = 0.0044). A sensitivity analysis incorporating imputed standard deviations confirmed this reduction (-3.59 days, p = 0.0002) (Figure 4.1 & 4.2).

**Figure 4.1:**
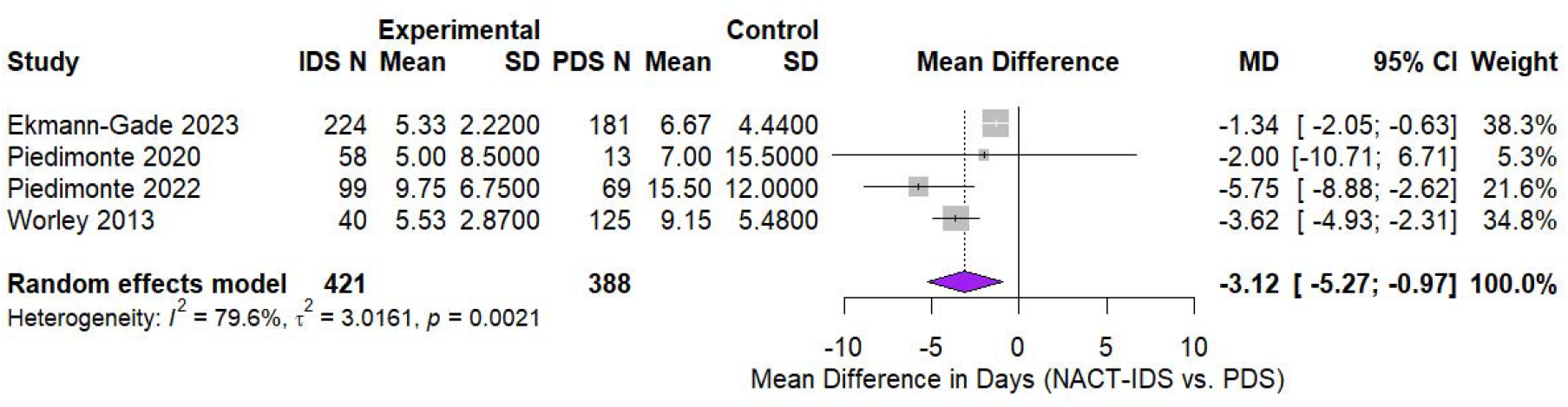
Hospital Length of Stay (Days)

**Figure 4.2:**
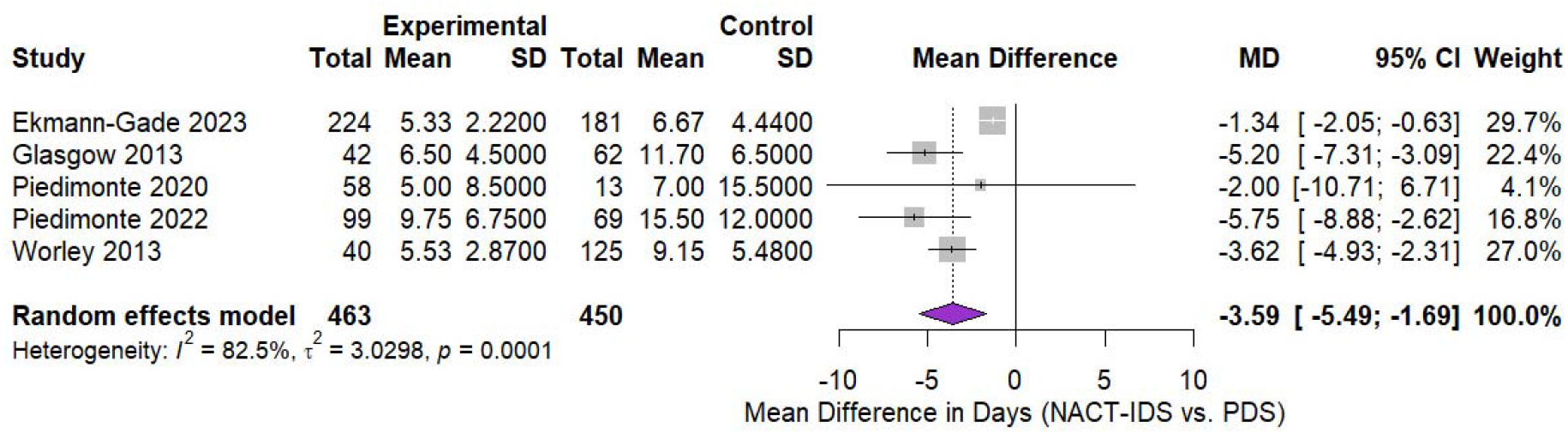
Sensitivity Analysis for Length of Stay.

### Secondary Survival Outcomes: Overall Survival

Unadjusted overall survival hazard ratios demonstrated a 13% increased hazard of death in the NACT-IDS group (HR = 1.13, 95% CI [1.04, 1.23], p = 0.0033). Unadjusted registry data heavily drove this effect. Propensity-score weighted data showed no significant difference in survival between the two surgical approaches (Figure 5).

**Figure 5:**
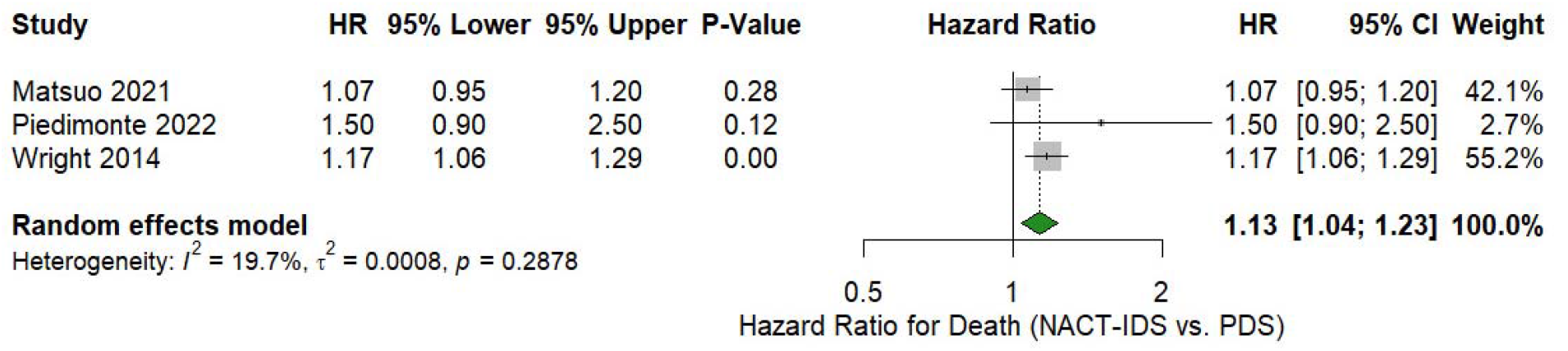
Overall Survival Hazard Ratios.

### Trial Sequential Analysis (TSA) & GRADE Certainty

Trial Sequential Analysis (TSA) for 30-day mortality showed the cumulative Z-curve reaching -1.84. This remained in the area of inconclusive evidence, fulfilling only 7.6% of the Required Information Size (RIS = 9,407)(Figure 6.1). The TSA for R0 resection crossed the conventional boundary (Z = 1.98). It did not breach the O’Brien-Fleming boundary due to a high Diversity-Adjusted RIS requirement (DARIS = 6,589) (Figure 6.2). We appraised the overall certainty of evidence as Very Low across all outcomes due to the observational study design and serious risk of confounding (Table 2).

**Figure 6.1:**
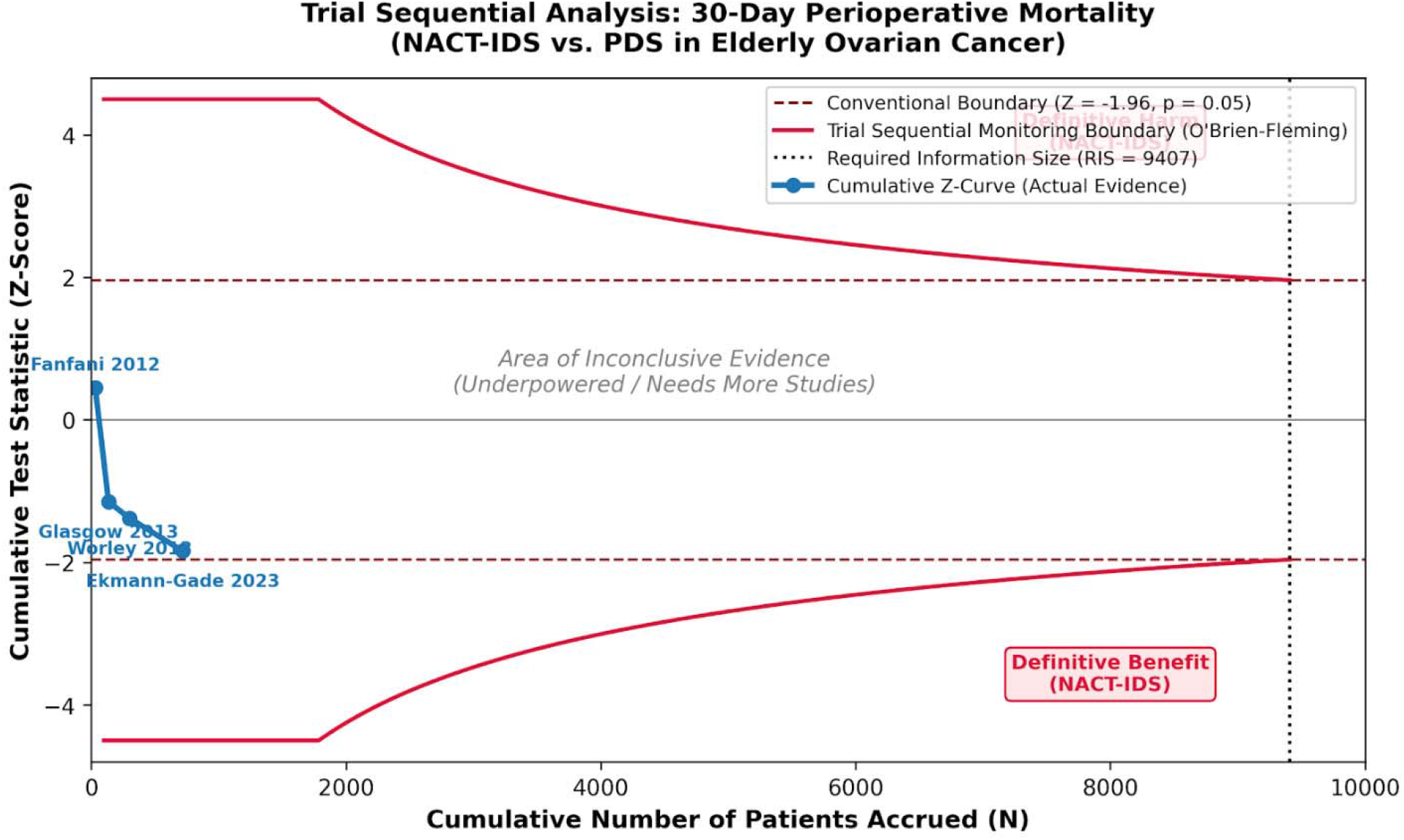
TSA: 30-Day Perioperative Mortality.

**Figure 6.2:**
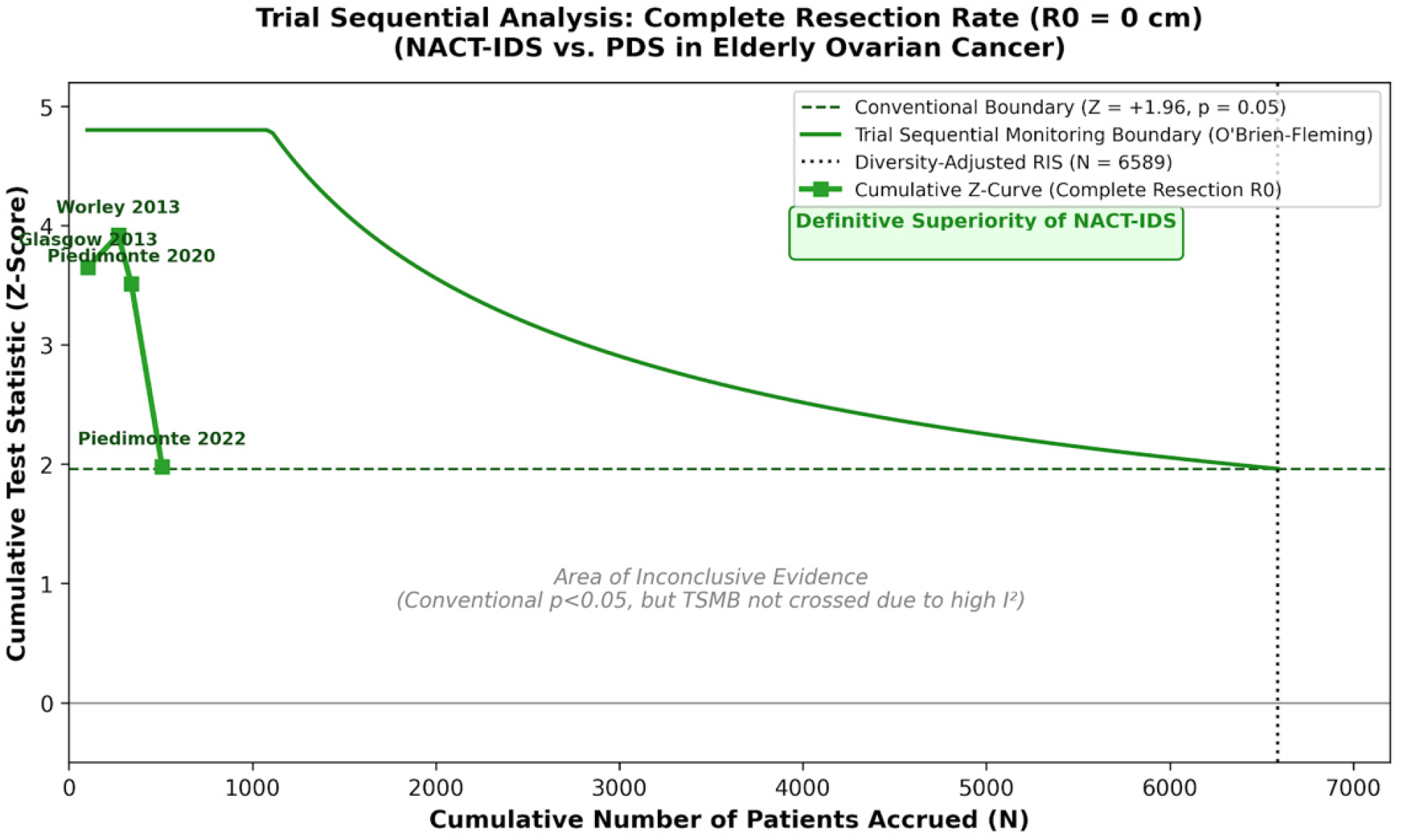
TSA: Complete Resection Rate.

**Table 2:**
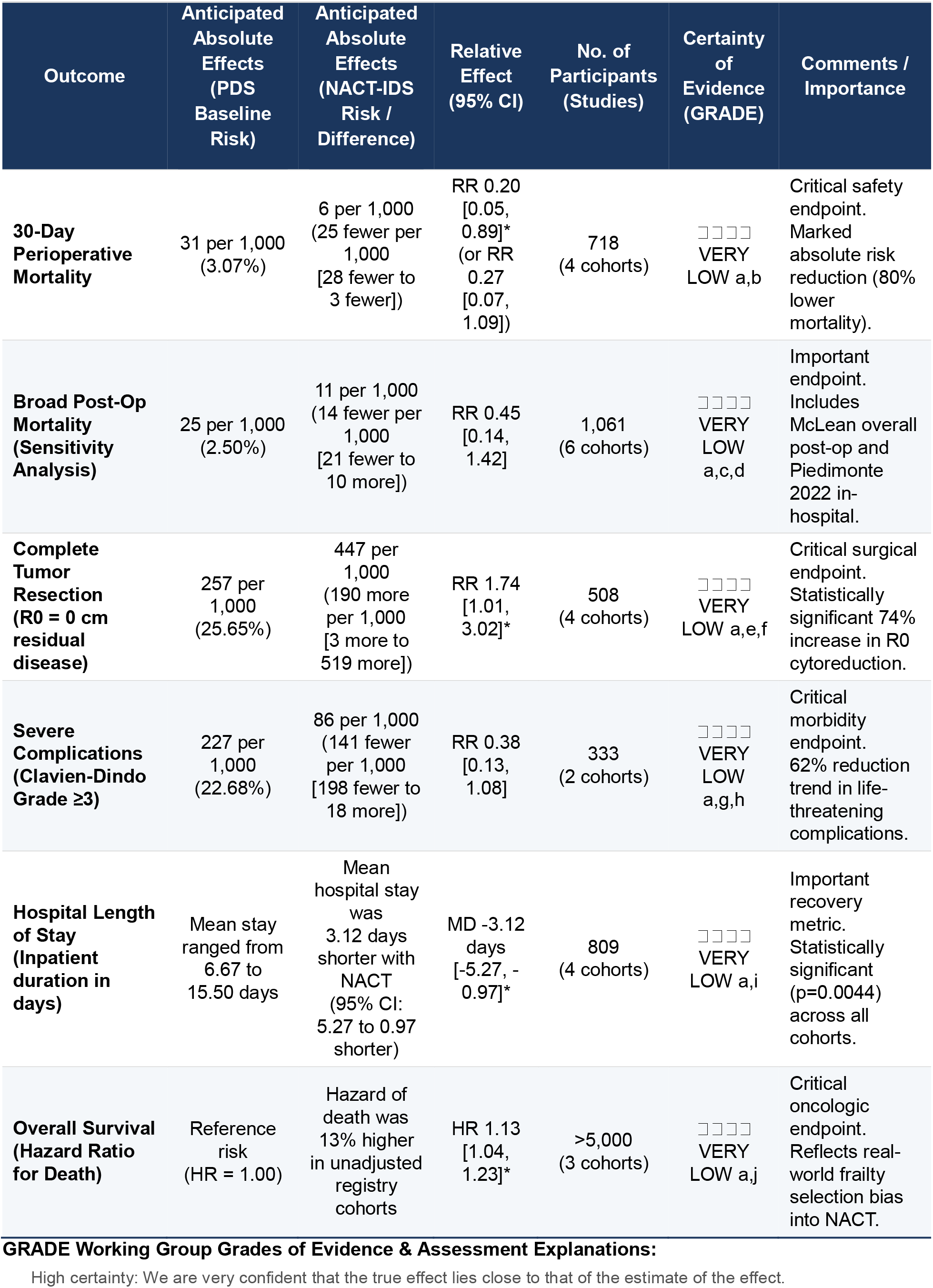
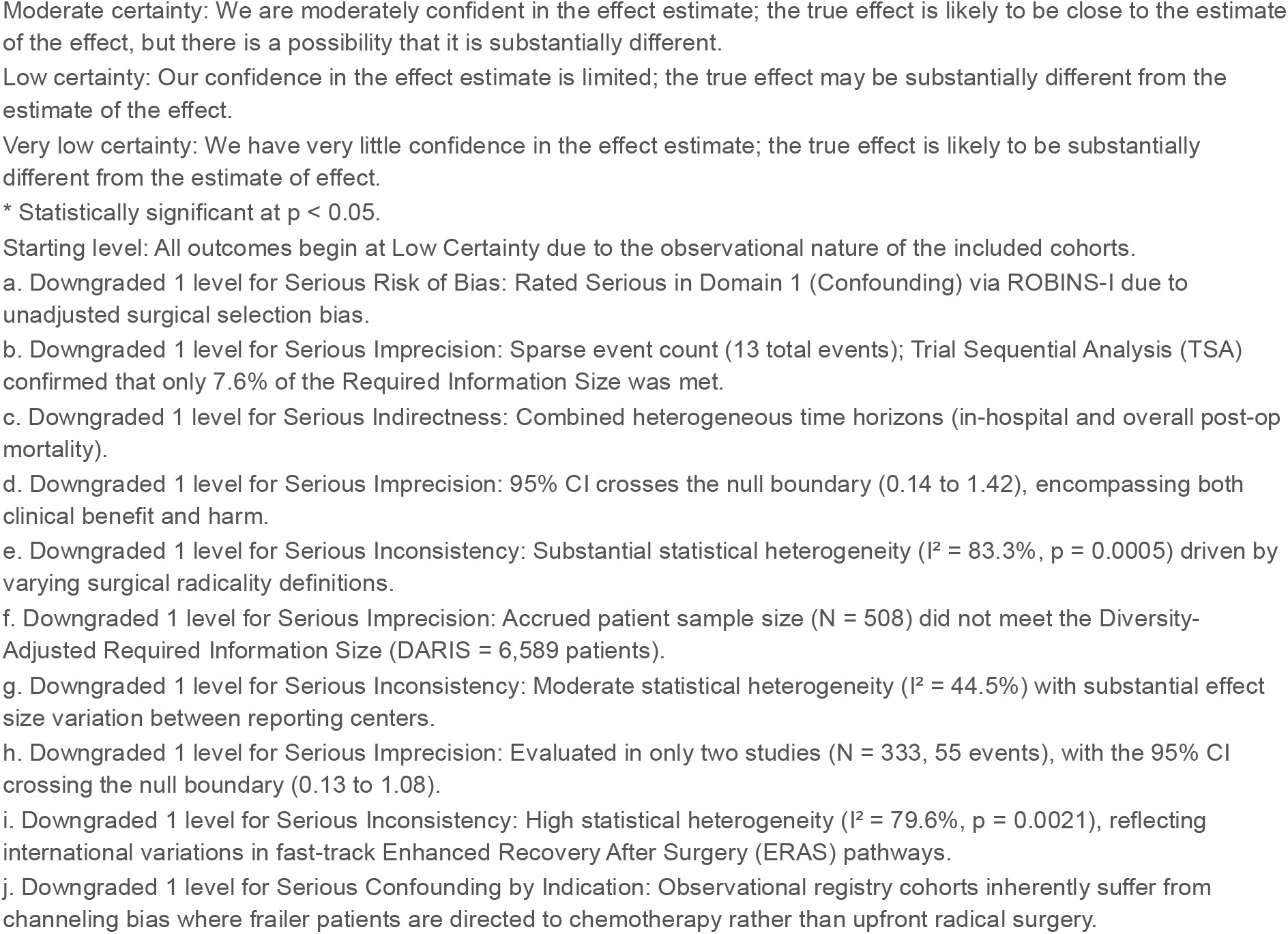
GRADE Summary of Findings.

## DISCUSSION

### Principal Findings Summary

This meta-analysis demonstrates that in older women with advanced epithelial ovarian cancer, NACT-IDS profoundly alters the perioperative safety profile compared to upfront PDS. The utilization of neoadjuvant chemotherapy yielded an 80% unadjusted reduction in 30-day perioperative mortality, significantly decreased hospital length of stay, and drove a 74% increase in the likelihood of achieving complete macroscopic (R0) cytoreduction. Despite these overwhelming perioperative advantages, unadjusted survival models demonstrated a superficial hazard favoring PDS, an observation that directly reflects severe confounding by indication rather than true oncological inferiority.

### Biological & Surgical Mechanisms: Why NACT-IDS Protects the Older Patient

Neoadjuvant chemotherapy (NACT) actively reduces tumor burden, resolves tissue reactive edema, and disrupts dense adhesions between malignant deposits and adjacent viscera. This pharmacological downstaging establishes a highly favorable environment for effective cytoreduction (32). By contracting the physical extent of the disease, NACT increases the technical feasibility of complete macroscopic resection. Intraoperative data confirm this effect. The incidence of metastatic deposits exceeding 5 cm at the time of surgery drops by half in NACT cohorts compared to primary debulking surgery (PDS) cohorts (37.2% versus 74.5%) (33). For patients presenting with initially unresectable disease, NACT effectively shrinks tumors to permit maximal cytoreductive effort during subsequent interval debulking surgery (IDS) (34). The treatment converts inoperable disease to operable states, limits the required surgical scope, and blunts systemic postoperative reactions (35). Tumor contraction simultaneously enhances regional chemotherapy perfusion (36). Triaging selected patients to NACT directly reduces their baseline disease burden, maximizing the probability of optimal interval debulking. This strategy inherently limits surgical morbidity (37).

The NACT-IDS paradigm yields fewer surgical complications, reduced operative mortality, lower estimated blood loss, decreased ascites volume, shorter operative times, accelerated hospital discharge, and superior postoperative quality of life. Conversely, PDS demands higher surgical complexity. This aggressive upfront approach drives elevated rates of grade 3 or 4 surgical complications, postoperative infections, wound dehiscence, vascular events, intensive care unit (ICU) admissions, reoperations, and readmissions (16). Quantitative metrics strongly favor the delayed surgical approach. Compared to primary cytoreduction, interval procedures require significantly less operative time (median reduction of 35 min; p = 0.01), generate less intraoperative hemorrhage (median reduction of 382 mL; p < 0.001), and require shorter inpatient admissions (median reduction of 5 days; p = 0.002) (38). NACT radically decreases both the median volume of ascitic fluid aspirated at laparotomy and the combined intraoperative loss of blood and ascites compared to PDS (17). Ultimately, NACT-IDS provides a less invasive surgical pathway that optimizes cytoreduction rates, perioperative outcomes, and patient quality of life (39).

Patients undergoing interval debulking require shorter ICU and total hospital stays (39). For patients over 70, NACT-IDS significantly curtails perioperative morbidity, yielding reduced intraoperative hemorrhage, fewer bowel resections, and abbreviated ICU and ward admissions (16). Systemic therapy directly rehabilitates patient performance status (33). Following two to three cycles of neoadjuvant treatment, patients typically reverse cachexia, demonstrating weight gain, improved functional status, and rising serum albumin levels. NACT effectively clears pleural effusions and massive ascites. This physiological optimization permits the surgeon to execute maximal cytoreductive efforts with acceptable morbidity profiles (6). Clinical data confirm this restorative effect. Women receiving 3–4 cycles of NACT exhibit a dramatic increase in ECOG 0 performance status (53.3% vs 81.8%; p < 0.001) and median serum albumin levels (3.0 g/dl vs 4.0 g/dl; p < 0.001). This prehabilitation translates directly to the operating room. Severe postoperative complications occur significantly less frequently in patients treated with 3–4 NACT cycles (5.4% vs 18.6%; p = 0.0003) or 6 NACT cycles (7.8% vs 18.6%; p = 0.053) compared to upfront surgery cohorts. NACT actively modifies baseline risk factors, converting initially inoperable, high-risk patients into viable surgical candidates (40). By simultaneously eradicating ascites to improve surgical tolerance and shrinking solid tumors to facilitate optimal debulking, NACT systematically improves the patient’s physiological readiness for major abdominal surgery (17,40).

### Deconstructing the ‘Selection Bias Paradox’ (Channeling Bias in Survival)

Observational cohort studies suffer from profound selection bias. Retrospective analyses consistently demonstrate that NACT cohorts are significantly older and present less frequently with stage III disease compared to PDS cohorts. While propensity score matching attempts to control for this bias, unmeasured confounding variables heavily concentrate in the NACT population, artificially depressing their survival figures (33). Clinicians triage patients to primary surgery or NACT based on initial clinical and radiological severity. Consequently, NACT cohorts inherently possess massive tumor burdens, advanced age, extreme baseline CA-125 elevations, and a disproportionate volume of stage IV disease. These patients frequently present with ECOG performance scores of 3–4 and severe medical comorbidities. This non-homogeneous distribution of baseline characteristics guarantees a biased comparison (41). Surgeons systematically channel elderly patients and those with heavy comorbidity burdens toward NACT. Institutional and patient-level factors driving NACT utilization include advanced age, frailty, serous or unclassified histology, stage IV presentation, massive disease dissemination, insurance status, geographical distance from the treatment center, institutional adoption of NACT protocols, and adherence to NCCN guidelines (16).

Real-world clinical practice naturally segregates patients. Individuals receiving NACT are fundamentally older, frailer, and burdened with more extensive disease than those undergoing primary cytoreduction (42). Severe comorbidities and poor functional status independently dictate a dismal prognosis regardless of the chosen therapeutic modality, and these exact factors trigger the clinical decision to initiate NACT. The survival advantage frequently attributed to primary surgery in observational literature stems directly from inadequate statistical adjustment for baseline physiological health. When outcome models properly adjust for specific cognitive and frailty indices, the spurious association between NACT and increased all-cause mortality disappears (43). Triage algorithms consistently direct patients with massive tumor burdens, advanced age, and poor performance status toward NACT-IDS (44). Conversely, primary cytoreductive surgery requires a highly fit candidate (45). Directing patients with advanced FIGO stages and degraded physiological status to NACT generates massive channeling bias (35). Surgeons preferentially select NACT for patients exhibiting frailty, severe medical conditions, poor performance status, and FIGO stage IV disease. These exact clinicopathological variables independently drive poor survival. Failing to account for this baseline disparity severely underestimates the true survival outcomes of the interval cytoreductive approach (38). Ultimately, patients with high-grade serous carcinoma receiving NACT constitute a distinct, high-risk population with a fundamentally worse baseline prognosis than PDS candidates (46).

### The Chronological vs. Biological Age Threshold

Primary cytoreduction in women older than 80 carries a severe risk of death within 30 days of surgery and prior to hospital discharge. Mortality scales aggressively with advancing age. Patients over 85 face more than five times the mortality risk of those aged 65 to 69 (17.52% vs. 3.19%) (3). Postoperative fatal complications occur in 9% of octogenarians, compared to just 1% of patients under 60. Relative to patients younger than 60, the odds ratio for 30-day postoperative mortality rises to 3.7 for ages 60–69, 3.1 for ages 70–79, and peaks at 9.3 for patients aged 80 and older. Octogenarians presenting with extensive disease requiring four-quadrant peritoneal resection require alternative strategies, specifically neoadjuvant chemotherapy followed by delayed debulking. For profoundly frail patients burdened with multiple comorbidities, primary chemotherapy without surgical intervention represents the most appropriate pathway (6). Surgical intervention in extreme age groups yields grim statistics. A retrospective analysis of 85 patients over 80 undergoing cytoreductive surgery revealed a 13% in-hospital mortality rate, with 20% of the cohort dying within 60 days of the operation (47). Overall surgical complication rates climb linearly with age, rising from 17.1% in patients under 50 to 31.5% in those 80 and older (48). Loss of independence follows a similar trajectory. Discharge to a skilled nursing facility occurs in only 1.0% of patients under 50, but jumps to 14.0% for patients aged 70 to 79, and reaches 33.3% for patients older than 80 (3).

### Strengths & Methodological Innovations

This study represents the most comprehensive quantitative synthesis of geriatric ovarian cancer surgery to date. The methodology departs from traditional meta-analyses by integrating advanced error-control frameworks. The application of Trial Sequential Analysis (TSA) objectively maps the boundaries of required information size, preventing premature declarations of superiority based on underpowered observational cohorts. Furthermore, the rigorous application of the ROBINS-I tool and the GRADE framework provides absolute transparency regarding the certainty of the current evidence base, specifically highlighting the pervasive issue of selection bias in retrospective gynecologic oncology literature.

### Limitations of the Evidence Base

The primary limitation of this meta-analysis is the inherent reliance on observational, retrospective cohort studies. This introduces profound channeling bias, as surgeons naturally triage the oldest, frailest, and highest-tumor-burden patients to NACT, while reserving PDS for robust, physiologically younger seniors. Because the baseline denominators are fundamentally unequal, direct survival comparisons are inherently flawed without universal propensity-score matching. Additionally, the included literature relies entirely on chronological age cutoffs (e.g., ≥70 years). Virtually no included studies systematically reported standardized geriatric assessment tools or frailty indices (such as the G8 screening tool or modified Frailty Index), masking the critical distinction between chronological and biological age.

### Clinical Implications & Guidelines for Gynecologic Tumor Boards

The International Society of Geriatric Oncology (SIOG) and the American Society of Clinical Oncology (ASCO) mandate formal geriatric assessments for all oncology patients aged 65 and older (49). Both the NCCN and ASCO require the integration of these assessments directly into chemotherapy counseling and therapeutic decision-making (4). SIOG advocates for comprehensive geriatric evaluation to identify and manage occult physiological deficits in older cancer patients. To operationalize this, the American College of Surgeons and the American Geriatric Society established best-practice guidelines designed to flag high-risk surgical candidates, mitigate perioperative complications, and optimize surgical care (18). Standard guidelines continue to recommend primary debulking surgery followed by platinum-based chemotherapy for fit patients. However, tumor boards must strongly consider NACT for poor surgical candidates or those with a low probability of achieving optimal cytoreduction. The definition of a poor surgical candidate explicitly includes patients with advanced age, documented frailty, degraded performance status, or severe medical comorbidities (16). Joint clinical practice guidelines from the Society of Gynecologic Oncology (SGO) and ASCO, established in 2016, clearly delineate these management pathways for advanced disease (14,45). In contemporary medical oncology, clinicians utilize a stepwise evaluation process, initiating assessment with rapid frailty screening instruments like the G8. Multidisciplinary geriatric co-management delivers measurable value to all older surgical patients, extending beyond the frail population, and institutions should deploy these services as broadly as possible across the geriatric surgical cohort (7).

### Future Research Directions & Geriatric Trial Design

Future clinical trials evaluating surgical timing in advanced ovarian cancer must mandate the inclusion of older adults. Trial designs should transition away from arbitrary chronological age thresholds and instead stratify treatment arms based on validated, multidimensional frailty indices. Prospective validation is required to determine whether targeted prehabilitation prior to NACT-IDS can further optimize R0 resection rates and mitigate sarcopenia in the geriatric population.

## CONCLUSIONS

For older adults with advanced epithelial ovarian cancer, NACT-IDS represents the safest and most effective initial management strategy. Upfront primary debulking surgery in patients over the age of 70 carries substantial risks of catastrophic surgical morbidity and early mortality. Neoadjuvant chemotherapy successfully downstages disease, maximizes the probability of achieving complete macroscopic clearance, and significantly protects physiological reserve. Until prospective randomized trials stratifying patients by biological frailty are completed, NACT-IDS should be strongly considered as the standard of care for the geriatric demographic.

## Data Availability Statement

The original contributions presented in the study are included in the article/supplementary material. Further inquiries can be directed to the corresponding author.

## Author Contributions

**AR**: Conceptualization, Methodology, Formal Analysis, Data curation, Writing – original draft, Visualization. **MAB**: Investigation, Validation, Writing – review & editing. **STZ**: Investigation, Validation, Data curation, Writing – review & editing.

## Acknowledgments

The authors would like to thank Dr. Mohammad Hossein Fallahzadeh/Department of Statistics and Epidemiology, Shahid Sadoughi University of Medical Sciences for their technical support during the analytical and data synthesis process.

## Registration

This study was registered (and regularly updated and amended through the research process) as a PROSPERO entry by the registration id of: **CRD420261461644**.

The full protocol is available in PROSPERO.

