## Supplementary figures and images for "Perioperative Safety, Surgical Morbidity, and Survival Outcomes of Neoadjuvant Chemotherapy (NACT-IDS) versus Primary Debulking Surgery (PDS) in Elderly Patients with Advanced Epithelial Ovarian Cancer: A Systematic Review and Meta-Analysis"

### supplimentary figure 3.1

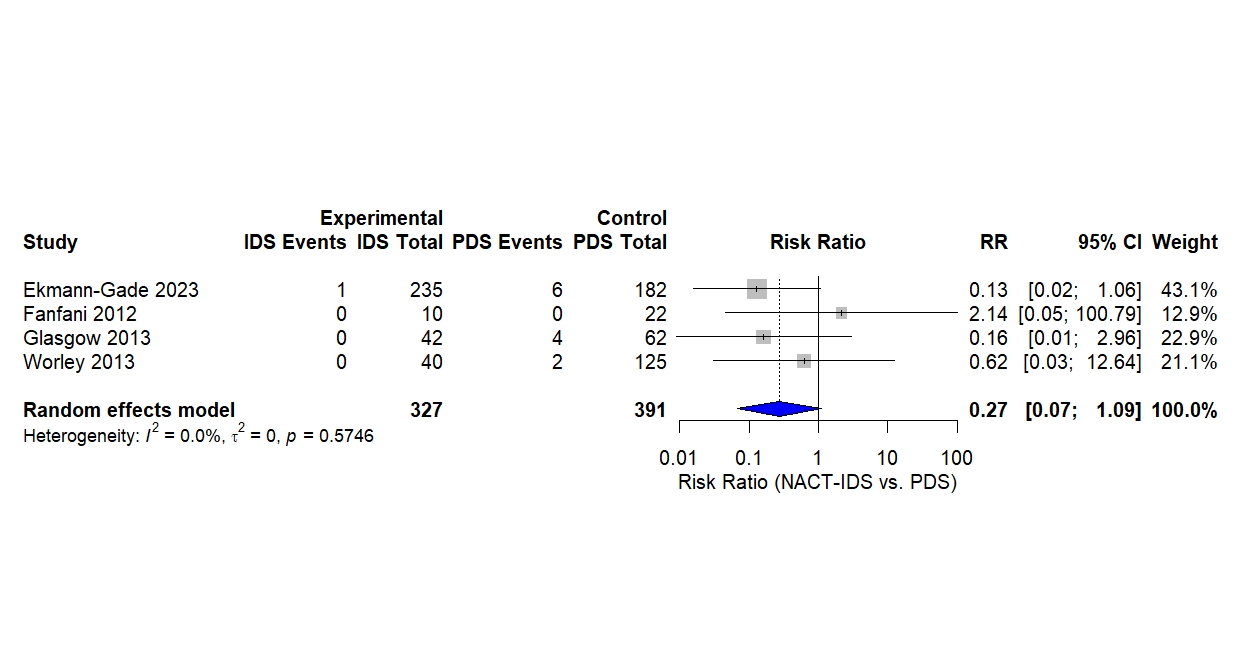

### supplimentary figure 3.2

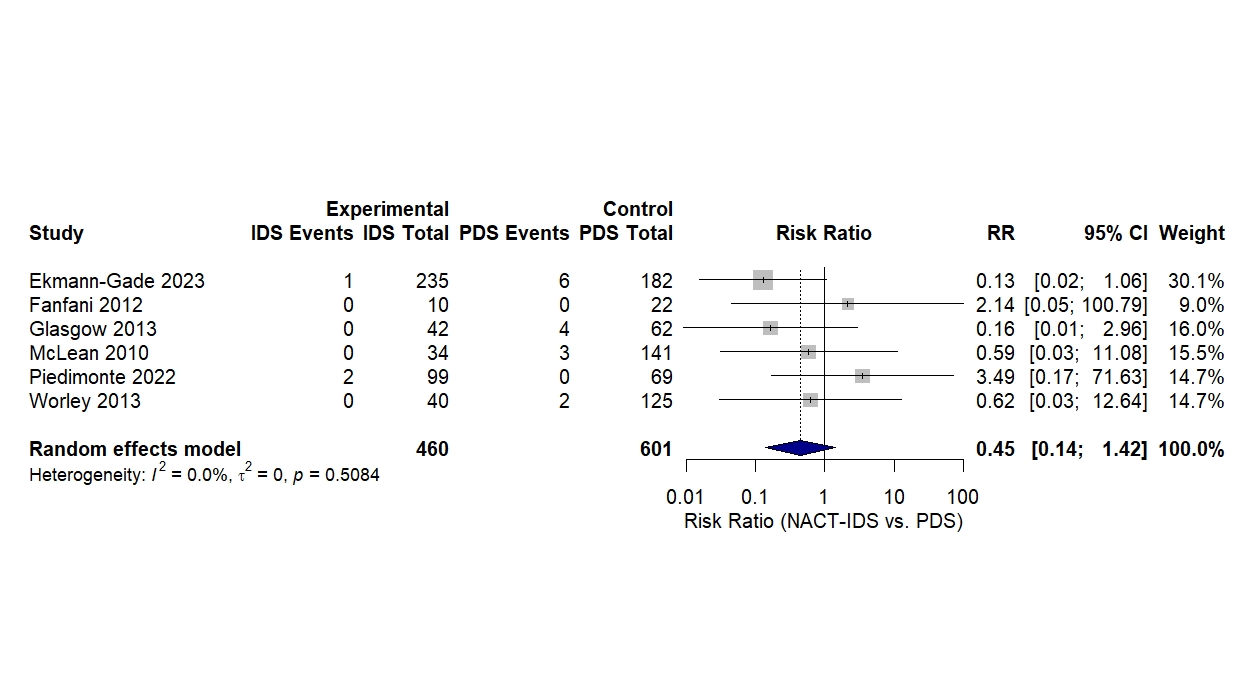
